# The interaction effect between hemoglobin and hypoxemia on COVID-19 mortality in a sample from Bogotá, Colombia: An exploratory study

**DOI:** 10.1101/2022.02.07.22270640

**Authors:** Andrés Felipe Patiño-Aldana, Angela María Ruíz-Sternberg, Angela María Pinzón-Rondón, Nicolás Molano-González, David Rene Rodríguez Lima

## Abstract

**Purpose:** We aimed to assess the effect of hemoglobin (Hb) concentration and oxygenation index on COVID-19 patients’ mortality risk.

**Patients and methods:** We retrospectively reviewed sociodemographic and clinical characteristics, laboratory findings, and clinical outcomes from patients admitted to a tertiary care hospital in Bogotá, Colombia. We assessed exploratory associations between oxygenation index and Hb concentration at admission and clinical outcomes. We used a generalized additive model (GAM) to evaluate the nonlinear relations observed and the classification and regression trees (CART) algorithm to assess the interaction effects found.

**Results:** From March to July 2020, 643 patients were admitted, of which 52% were male. The median age was 60 years old, and the most frequent comorbidity was hypertension (35.76%). The median value of SpO_2_/FiO_2_ was 419, and the median Hb concentration was 14.8 g/dL. The mortality was 19.1% (123 patients). Age, sex, and history of hypertension were independently associated with mortality. We described a nonlinear relationship between SpO_2_/FiO_2_, Hb concentration and neutrophil-to-lymphocyte ratio with mortality and an interaction effect between SpO_2_/FiO_2_ and Hb concentration. Patients with a similar oxygenation index had different mortality likelihoods based upon their Hb at admission. CART showed that patients with SpO_2_/FiO_2_ < 324, who were older than 62 years, and had an Hb of ≥ 16 g/dl had the highest mortality risk (96%). Additionally, patients with SpO_2_/FiO_2_ > 324 but Hb of < 12 and neutrophil-to-lymphocyte ratio of > 4 had a higher mortality likelihood (57%). In contrast, patients with SpO_2_/FiO_2_ > 324 and Hb of > 12 g/dl had the lowest mortality risk (10%).

**Conclusion:** We found that a decreased SpO_2_/FiO_2_ increased mortality risk. Extreme values of Hb, either low or high, showed an increase in likelihood of mortality. However, Hb concentration modified the SpO_2_/FiO_2_ effect on mortality; the likelihood of death in patients with low SpO_2_/FiO_2_ increased as Hb increased.

## Introduction

On December 31, 2019, SARS-CoV-2 emerged in China and rapidly spread worldwide, with more than 293 million infections and 5.4 million deaths.[1] In Colombia, which is a developing country, as of January 5th, there have been more than 5.2 million cases, with more than 130,000 deaths in the national territory. In Bogota, the most affected city in the country, there have been more than 1.49 million people infected and 27,846 people have died.[2,3]

Several studies worldwide have associated factors with mortality in COVID-19 patients. Most studies have been consistent with risk factors for severe disease. Studies in Asia reported age, history of smoking, vital signs at admission, albumin, C-reactive protein, and proinflammatory cytokines as risk factors for disease progression.[4,5] Studies in the USA described factors such as oxygenation (SpO_2_/FiO_2_) at admission, age, heart failure, sex, nursing home residency, respiratory rate, and body mass index (BMI) as the main predictors associated with COVID-19 critical illness.[6,7] These factors might be linked to a hyperinflammatory syndrome, which favors an exaggerated immune response and organ failure when facing a viral infection.[8,9]

Some ecological studies have evaluated COVID-19 mortality in high-altitude places, but the results are contradictory. Some authors reported higher mortality in men younger than 65 years old living in the USA and Mexico at >2,000 m elevation than in those located <1,500 m.[10] Other studies developed in an Andean population (Colombia and Perú) showed a negative correlation between high altitude and COVID-19 mortality and lower mortality excess.[11–13] People who permanently live in high-altitude places develop adaptative mechanisms against low atmospheric oxygen pressure exposure. In locations such as Bogotá, which has an altitude of more than 2,500 meters, there is a decrease in mean SpO_2_ values among its population.[14] People who live in high altitude locations develop ventilatory, cardiovascular, reproductive, and even cognitive changes. [15,16]

As part of the compensatory mechanisms, erythrocyte concentration in the blood increases. Initially, it is caused by a fast reduction in plasma volume. Additionally, hypoxia leads to an erythropoietic stimulus through hypoxia-inducible factor 1-alpha (HIF1-α) activation that binds hypoxia-inducible factor 1-beta (HIF1-β). Together, they act as a promoting factor for erythropoietin (EPO) stimulating red blood cell (RBC) production.[17] High altitude also leads to hyperventilation driven by an increased tidal volume and respiratory rate. Living in a high-altitude place is related to alveolar and arterial hypoxemia. A person at sea level will have an alveolar oxygen pressure between 90 and 100 mm Hg, a resident at Bogotá will reach a PaO2 between 60 and 70 mmHg, while someone at 5,100 m elevation will have a PaO2 of 43 mmHg.[18–20] The adaptative mechanisms allow for the survival of populations conditioned to these harsh environments. However, studies of people living at high altitudes have described increases in the O_2_ maximum consumption as they descend to sea level, highlighting the physiologically challenging condition imposed by low oxygen pressures for oxygen uptake and utilization.[18]

Numerous studies have proposed hemoglobin (Hb) as a biomarker of severe COVID-19.[21] Anemia is a marker of chronic disease and a risk factor in critical ill patients. Limited oxygen-carrying capacity and delivery might play a crucial role in organ failure development. Thus, increasing the likelihood of severity and mortality.[21,22] A recent meta-analysis found a lower mean Hb concentration between moderate and severe cases, but did not find significant differences between survivors and nonsurvivors.[21]

Adaptative mechanisms to high altitude are related to oxygen intake, transport, and the availability for tissues. Patients with acute respiratory infections who live in high altitude face a convergence between pathological and adaptive processes. Mechanisms that could be interrelated moderating the disease outcome. Our research aimed to assess the effect of Hb and oxygenation index on COVID-19 mortality.

## Materials and methods

### Study population

We performed an observational, retrospective study between March and July 2020. We followed a cohort of COVID-19 patients admitted to the Hospital Universitario Mayor Méderi (HUM) in Bogotá, Colombia. In this study, we used a cutoff date of July 2020. We included patients with confirmed infection of SARS-CoV-2 by RT–PCR or antigen. We excluded patients with a history of anemic, lymphoproliferative, or myelodysplastic syndromes. We also excluded patients with information of permanent residency under 2,500 meters above sea level (MASL) and without complete blood count (CBC) measurement. The Ethics Committee of the Universidad del Rosario approved the research protocol. Qualified professionals carried out all activities and procedures following the principles outlined in the Declaration of Helsinki.

### Variables and main outcomes

Data were collected from clinical records through a comprehensive review of hospital admission information. A CBC was taken only in patients with hospital admission criteria that required laboratory assessments, and who received at least a few hours of in-hospital observation. The CBC was processed in HUM’s clinical laboratory using a Sysmex XN-1000 hematology analyzer. In cases of low-flow systems that could not deliver constant FiO_2_, we took clinically recorded FiO_2_. A nasal cannula increased FiO_2_ by approximately 0.4% per liter/min. A nonrebreather mask with a flow output higher than 10 L/min was taken as 100% FiO_2_.[23] We calculated variables such as SpO_2_/FiO_2_, mean arterial pressure (MAP), and neutrophil to lymphocyte ratio (NLR). We defined the discharge condition (alive or dead) as the primary outcome.

### Statistical analysis

We reported qualitative variables as frequencies and percentages, and quantitative variables as the means and standard deviations or medians and interquartile ranges, depending on the normality of the distribution (tested by Shapiro–Wilk test). To assess the possible associations with mortality, we performed an exploratory analysis using the Mann–Whitney test or the chi-square of independence test for quantitative and qualitative variables, respectively. We made a purposeful selection of covariates for the multivariate model, as described by Hosmer, Lemeshow, and Sturdivant.[24] Appraising to nonlinear relations found, we built a generalized additive model (GAM), estimating linear and nonlinear effects. Instead of a single regression coefficient, GAM estimates a k number of base functions, which in sum describes the functional form of the relationship between dependent and independent variables.[25] We tested for interaction effects using tensor products in the GAM model. Notably, the tensor smoothing function (TI) calculates different k base functions for each variable included in the interaction effect, and it is useful for variables with different scales. [24,25]

In a second approach, we used the classification and regression tree (CART) algorithm to find the most relevant variables associated with the clinical outcome.[26] We set the overall significance level at 5%. The sampling method was not probabilistic and consecutive. We defined the sample size by convenience. Thus, the analyses were exploratory. We used R version 4.0.2 software for statistical analyses.

## Results

From March to July 2020, 1,000 patients were diagnosed with SARS-CoV-2 infection at HUM, and 650 met the inclusion criteria with in-hospital admittance and CBC measurement. We excluded seven patients because they met the exclusion criteria. Four of them had a history of anemia, two hematologic malignancies, and one permanent residency at low altitude (< 2,500 MASL), yielding 643 patients included in this study.

The median (interquartile range) age was 60 years old (46-73), and 52.56% of patients were male. The more frequent comorbidities were hypertension (35.76%), diabetes (18.04%), COPD (9.92%), and chronic kidney disease (CKD) (5.9%). The median MAP and SpO_2_/FiO_2_ were 92 mmHg (83.7-101.9) and 419 (359.5-438), respectively. Regarding the CBC on admission, the median white blood cell count (WBC) was 7.11 (5.29-9,71) × 10^3^ cells/µL, and the median concentration of Hb was 14.8 g/dL (13.5-15.9). Among the total sample, 123 patients (19.12%) died and 520 (80.87%) survived.

Table 1 shows the proportion of characteristics and median differences in vital signs and CBC by discharge condition with an exploratory hypothesis test result. We found a statistically significant difference in the proportion of males between survivors and nonsurvivors. Additionally, there was statistically significant difference in age and the number of patients with a history of hypertension, COPD, and CKD. Concerning vital signs, we found statistically significant differences in the median respiratory rate, FiO_2_, SpO_2_, and SpO_2_/FiO_2_. Regarding CBC, there were differences between survivors and nonsurvivors patients in terms of median WBC, RBC count, absolute lymphocyte count (ALC), absolute neutrophil count (ANC), Hb, NLR, and several percent white blood cell relations.

**Table 1.**
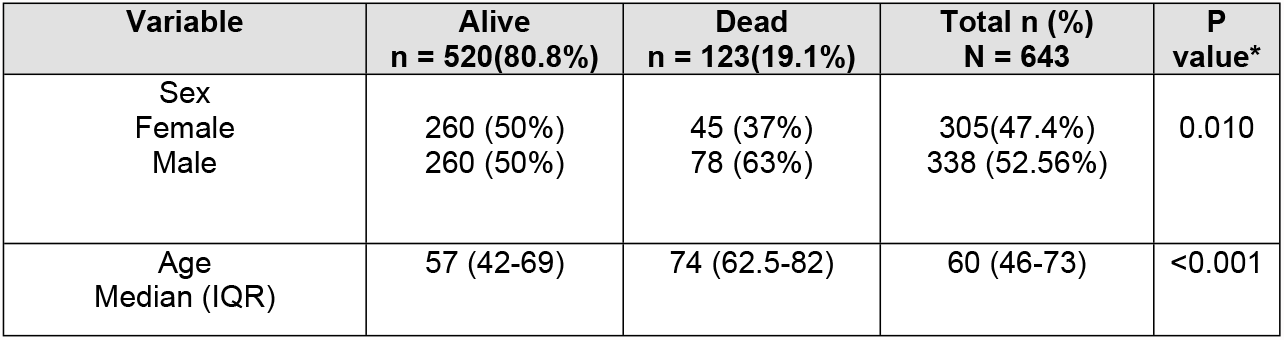
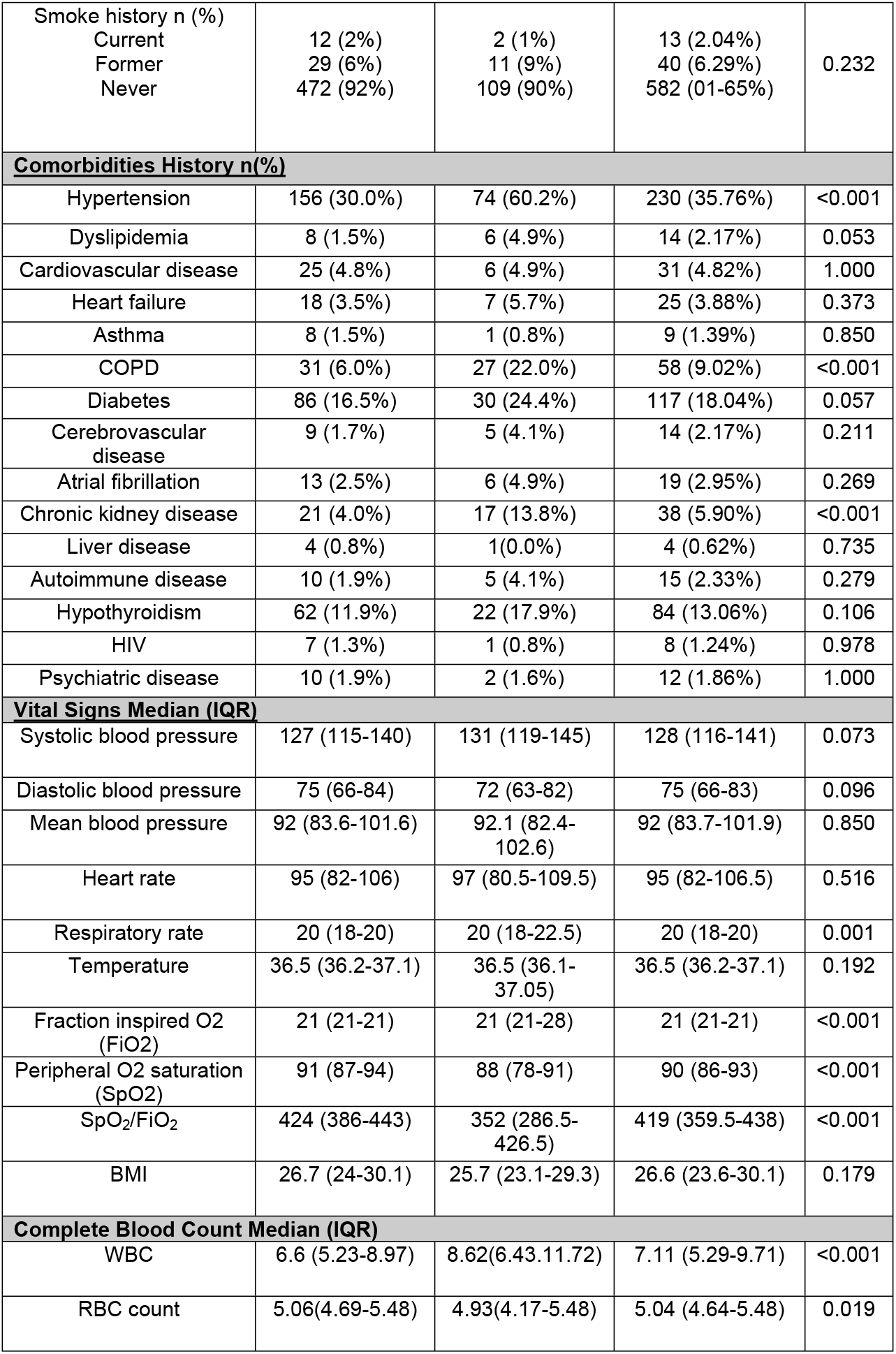
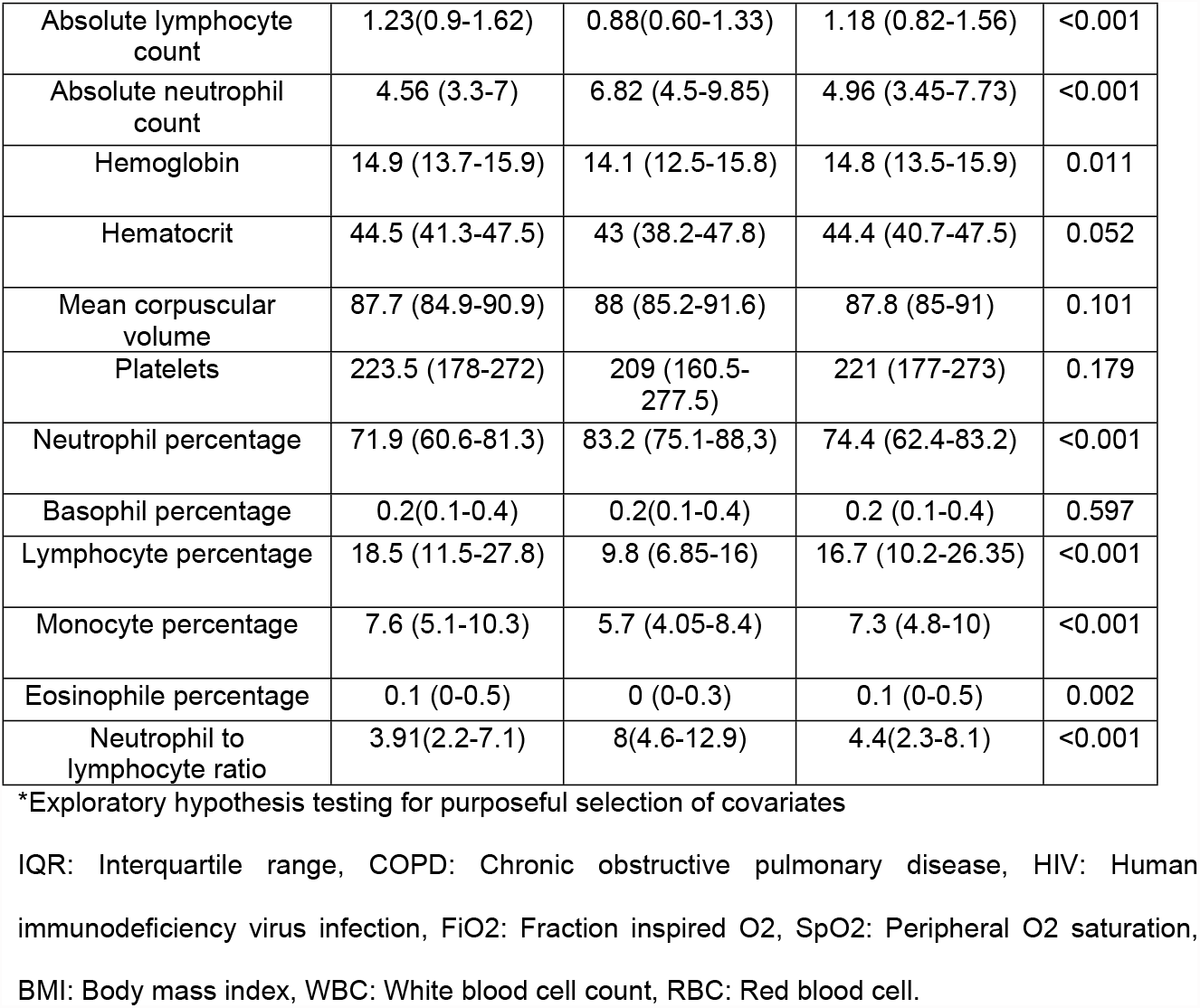
Demographic characteristics, comorbidity history, vital signs, and complete blood count by discharge condition in COVID-19 patients.

| Variable | Alive<br>n = 520(80.8%) | Dead<br>n = 123(19.1%) | Total n (%)<br>N = 643 | P<br>value* |
| --- | --- | --- | --- | --- |
| Sex |  |  |  |  |
| Female | 260 (50%) | 45 (37%) | 305(47.4%) | 0.010 |
| Male | 260 (50%) | 78 (63%) | 338 (52.56%) |  |
| Age |  |  |  |  |
| Median (IQR) | 57 (42-69) | 74 (62.5-82) | 60 (46-73) | <0.001 |
| Smoke history n (%) |  |  |  |  |
| Current | 12 (2%) | 2 (1%) | 13 (2.04%) | 0.232 |
| Former | 29 (6%) | 11 (9%) | 40 (6.29%) |  |
| Never | 472 (92%) | 109 (90%) | 582 (91.65%) |  |
| **Comorbidities History n(%)** |  |  |  |  |
| Hypertension | 156 (30.0%) | 74 (60.2%) | 230 (35.76%) | <0.001 |
| Dyslipidemia | 8 (1.5%) | 6 (4.9%) | 14 (2.17%) | 0.053 |
| Cardiovascular disease | 25 (4.8%) | 6 (4.9%) | 31 (4.82%) | 1.000 |
| Heart failure | 18 (3.5%) | 7 (5.7%) | 25 (3.88%) | 0.373 |
| Asthma | 8 (1.5%) | 1 (0.8%) | 9 (1.39%) | 0.850 |
| COPD | 31 (6.0%) | 27 (22.0%) | 58 (9.02%) | <0.001 |
| Diabetes | 86 (16.5%) | 30 (24.4%) | 117 (18.04%) | 0.057 |
| Cerebrovascular disease | 9 (1.7%) | 5 (4.1%) | 14 (2.17%) | 0.211 |
| Atrial fibrillation | 13 (2.5%) | 6 (4.9%) | 19 (2.95%) | 0.269 |
| Chronic kidney disease | 21 (4.0%) | 17 (13.8%) | 38 (5.90%) | <0.001 |
| Liver disease | 4 (0.8%) | 1(0.0%) | 4 (0.62%) | 0.735 |
| Autoimmune disease | 10 (1.9%) | 5 (4.1%) | 15 (2.33%) | 0.279 |
| Hypothyroidism | 62 (11.9%) | 22 (17.9%) | 84 (13.06%) | 0.106 |
| HIV | 7 (1.3%) | 1 (0.8%) | 8 (1.24%) | 0.978 |
| Psychiatric disease | 10 (1.9%) | 2 (1.6%) | 12 (1.86%) | 1.000 |
| **Vital Signs Median (IQR)** |  |  |  |  |
| Systolic blood pressure | 127 (115-140) | 131 (119-145) | 128 (116-141) | 0.073 |
| Diastolic blood pressure | 75 (66-84) | 72 (63-82) | 75 (66-83) | 0.096 |
| Mean blood pressure | 92 (83.6-101.6) | 92.1 (82.4-102.6) | 92 (83.7-101.9) | 0.850 |
| Heart rate | 95 (82-106) | 97 (80.5-109.5) | 95 (82-106.5) | 0.516 |
| Respiratory rate | 20 (18-20) | 20 (18-22.5) | 20 (18-20) | 0.001 |
| Temperature | 36.5 (36.2-37.1) | 36.5 (36.1-37.05) | 36.5 (36.2-37.1) | 0.192 |
| Fraction inspired O2 (FiO2) | 21 (21-21) | 21 (21-28) | 21 (21-21) | <0.001 |
| Peripheral O2 saturation (SpO2) | 91 (87-94) | 88 (78-91) | 90 (86-93) | <0.001 |
| SpO2/FiO2 | 424 (386-443) | 352 (286.5-426.5) | 419 (359.5-438) | <0.001 |
| BMI | 26.7 (24-30.1) | 25.7 (23.1-29.3) | 26.6 (23.6-30.1) | 0.179 |
| **Complete Blood Count Median (IQR)** |  |  |  |  |
| WBC | 6.6 (5.23-8.97) | 8.62(6.43-11.72) | 7.11 (5.29-9.71) | <0.001 |
| RBC count | 5.06(4.69-5.48) | 4.93(4.17-5.48) | 5.04 (4.64-5.48) | 0.019 |
| Absolute lymphocyte count | 1.23(0.9-1.62) | 0.88(0.60-1.33) | 1.18 (0.82-1.56) | <0.001 |
| Absolute neutrophil count | 4.56 (3.3-7) | 6.82 (4.5-9.85) | 4.96 (3.45-7.73) | <0.001 |
| Hemoglobin | 14.9 (13.7-15.9) | 14.1 (12.5-15.8) | 14.8 (13.5-15.9) | 0.011 |
| Hematocrit | 44.5 (41.3-47.5) | 43 (38.2-47.8) | 44.4 (40.7-47.5) | 0.052 |
| Mean corpuscular volume | 87.7 (84.9-90.9) | 88 (85.2-91.6) | 87.8 (85-91) | 0.101 |
| Platelets | 223.5 (178-272) | 209 (160.5-277.5) | 221 (177-273) | 0.179 |
| Neutrophil percentage | 71.9 (60.6-81.3) | 83.2 (75.1-88,3) | 74.4 (62.4-83.2) | <0.001 |
| Basophil percentage | 0.2(0.1-0.4) | 0.2(0.1-0.4) | 0.2 (0.1-0.4) | 0.597 |
| Lymphocyte percentage | 18.5 (11.5-27.8) | 9.8 (6.85-16) | 16.7 (10.2-26.35) | <0.001 |
| Monocyte percentage | 7.6 (5.1-10.3) | 5.7 (4.05-8.4) | 7.3 (4.8-10) | <0.001 |
| Eosinophile percentage | 0.1 (0-0.5) | 0 (0-0.3) | 0.1 (0-0.5) | 0.002 |
| Neutrophil to lymphocyte ratio | 3.91(2.2-7.1) | 8(4.6-12.9) | 4.4(2.3-8.1) | <0.001 |

\*Exploratory hypothesis testing for purposeful selection of covariates
IQR: Interquartile range, COPD: Chronic obstructive pulmonary disease, HIV: Human immunodeficiency virus infection, FiO2: Fraction inspired O2, SpO2: Peripheral O2 saturation, BMI: Body mass index, WBC: White blood cell count, RBC: Red blood cell.

We carried out a purposeful selection of covariates for modeling based on statistically significant differences found.[24] We found that age, sex, history of hypertension, SpO2/FiO2, Hb, and NLR were independently associated with in-hospital mortality. The relationship between age and mortality was linear. However, the association between SpO2/FiO2, Hb, and NLR and mortality was nonlinear. Thus, as stated earlier, we performed a GAM that included linear and smoothened terms. Moreover, we found a significant interaction term between SpO2/FiO2 and hemoglobin. Table 2 shows the effects of the linear terms in the independent and interaction GAM. These estimations were the same for both models. Table 3 shows the smoothed variables in both models and the significance of its smoothed function.

**Table 2.** Linear terms of independent effects and interaction generalized additive model

| Linear terms | Coefficient | P value | OR | CI 95% |
| --- | --- | --- | --- | --- |
| Intercept | -2.661 | <0.001 |  |  |
| Age | 0.044 | <0.001 | 1.05 | 1.03-1.06 |
| Sex: Male | 0.783 | 0.003 | 2.19 | 1.28–3.73 |
| Hypertension: Yes | 0.611 | 0.017 | 1.84 | 1.11–3.05 |

The estimated effects for linear terms, age, sex, and hypertension history were the same among the independent effects model and interaction model.

**Table 3.** Smoothed terms of independent effects and interaction generalized additive model

| Independent effects model |  | Interaction model |  |
| --- | --- | --- | --- |
| Smoothed terms | P Value<sup>a</sup> | Smoothed terms | P Value |
| S<sup>b</sup>(Neutrophil-to-lymphocyte ratio) | <0.001 | S(Neutrophil-to-lymphocyte ratio) | <0.001 |
| S (SpO<sub>2</sub>/FiO<sub>2</sub>) | <0.001 | TI<sup>c</sup>(SpO<sub>2</sub>/FiO<sub>2</sub>, Hemoglobin) | 0.011 |
| S (Hemoglobin) | <0.001 |  |  |
| R<sup>2</sup> = 0.3, AIC = 470 |  | R<sup>2</sup> = 0.252, AIC= 495 |  |

<sup>a</sup>P value shown with smoothed and tensor terms tested the H<sub>0</sub>: f(x) = 0. <sup>b</sup>S(): Smoothed term;
<sup>c</sup>TI(): Tensor interaction term.

It is noteworthy that the model with the interaction term had a higher Akaike information criterion (AIC) and a lower determination coefficient (R^2^) than the principal effects model, which came at the cost of higher complexity. Smoothed variables and interaction effects from the GAM model can be better understood visually. Figure 1 presents the functional form of the smoothened variable’s influence on mortality. Death probability had a positive correlation with NLR. Values of SpO_2_/FiO_2_ present a sharp increase in the mortality risk under 350-320. Hemoglobin at admission had a U-shaped correlation with mortality. Patients with low or high hemoglobin concentrations were more prone to die during hospitalization. However, the significant interaction term showed us that the Hb and SpO_2_/FiO_2_ effects on mortality were modified by each other.

**Fig. 1.**
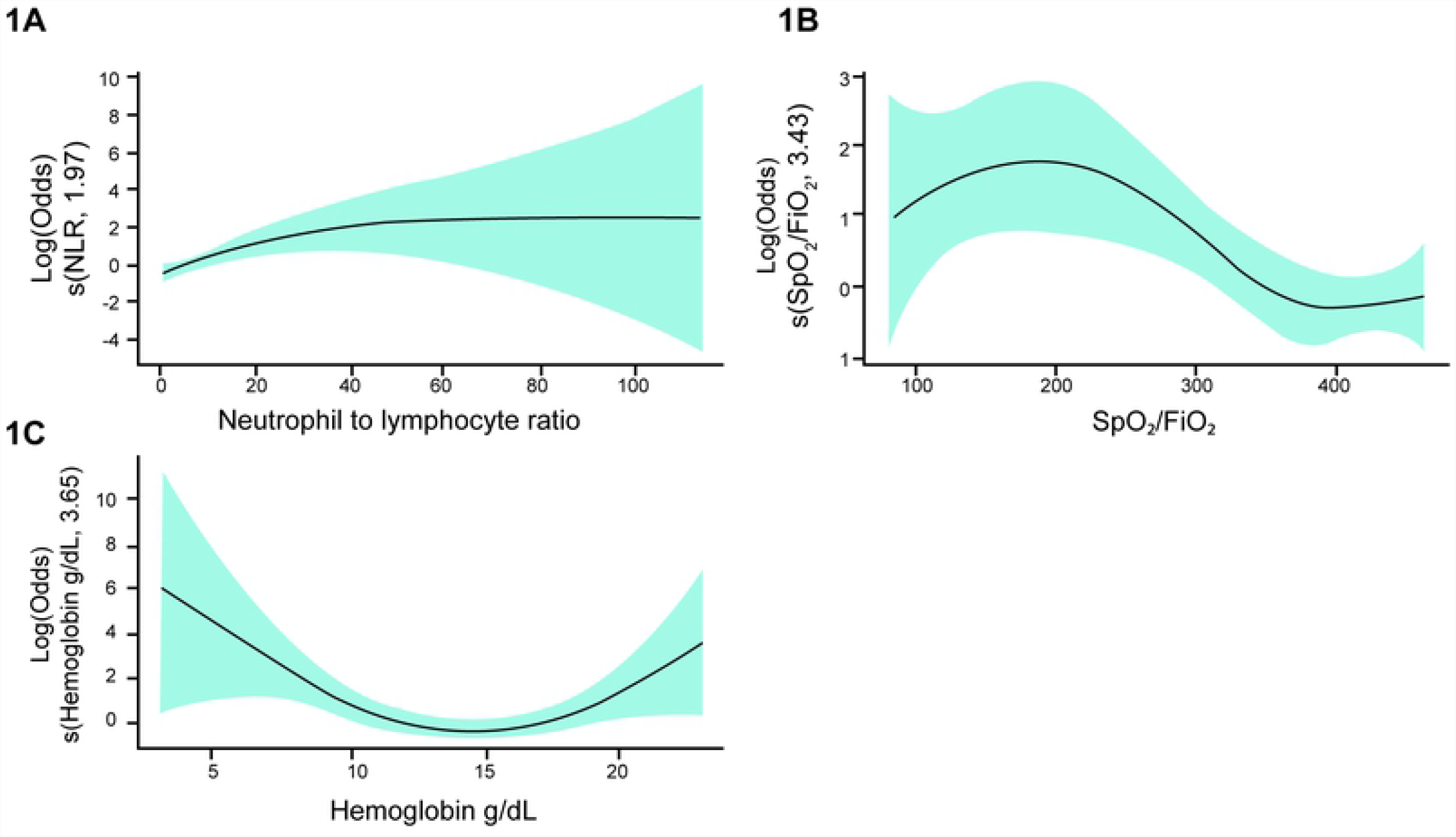
Partial effects of smoothed terms on mortality in GAM. Figure 1 shows the functional form of the partial effects on mortality of the smoothed terms in GAM. 1A) Positive correlation between NLR and mortality risk. 1B) Decreases in SpO_2_/FiO_2_ values lead to a sharp increase in death risk. 1C) U-shaped relationship between Hb concentration and mortality risk, patients with Hb either high or low values had an increased mortality risk. Inside parentheses are the smoothed term and its effective degrees of freedom (EDF), a summary statistic that reflects the degree of nonlinearity. NLR: Neutrophil to lymphocyte ratio, GAM: Generalized additive model.

The contour plot in Figure 2 shows how Hb modifies the SpO_2_/FiO_2_ effect on mortality. Notice how patients with similar SpO_2_/FiO_2_ at admittance had a different likelihood of death according to their Hb value at admission. For instance, death probability remained constant (10%) across all ranges of SpO_2_/FiO_2_ values for patients with an Hb between 12-15 g/dL. The likelihood of death was also 10% for those with a high Hb but SpO_2_/FiO_2_ >400. However, patients with SpO_2_/FiO_2_ <300 and Hb >15 g/dL had a 20% likelihood of death. This probability increased proportionally as the Hb concentration rose. It increased to 80% of death likelihood for patients with SpO_2_/FiO_2_ of 300 and Hb >20 g/dL and 90% for patients with SpO_2_/FiO_2_ <150 and an Hb of approximately 18 g/dL. Additionally, in patients with SpO_2_/FiO_2_ of 400 and Hb <10 g/dL, the death probability increased proportionally as Hb decreased, increasing up to 70 to 90% for patients with an Hb of approximately 5 g/dL. We further explored this interaction effect with a second approach, which is explained below.

**Fig. 2.**
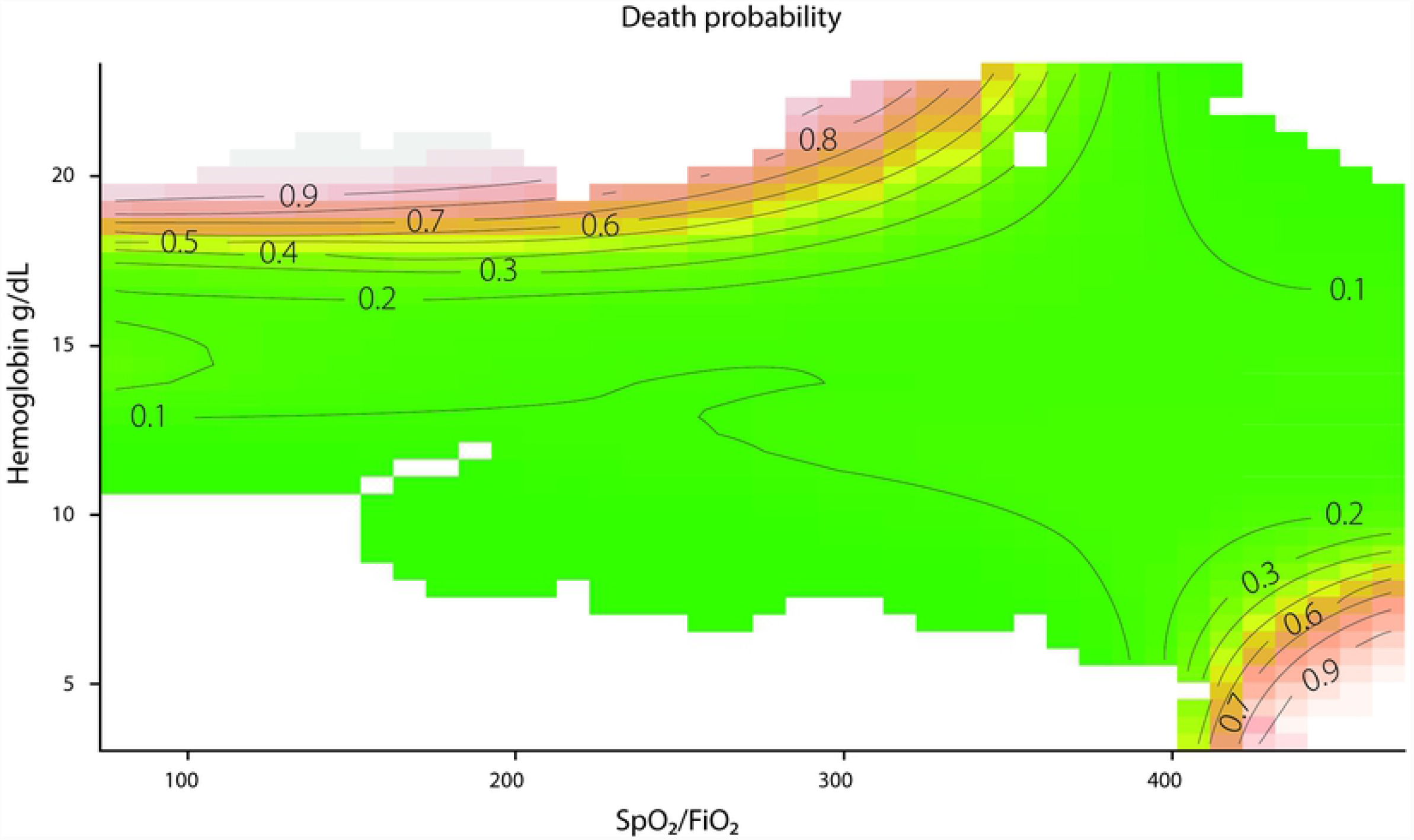
Interaction effect between SpO_2_/FiO_2_ and Hb on mortality. Figure 2. shows a contour plot with estimated probabilities of death for given SpO2/FiO2 values at different hemoglobin concentrations. Curves show areas where the risk of death is similar. Higher likelihoods are shown in red, while green indicates lower likelihoods. Patients with similar SpO2/FiO2 values had different probabilities of death according to hemoglobin concentration. Notice how decreases in SpO2/FiO2 get worse as hemoglobin concentration rise. Blanks in the plot represent data that were not observed.

Figure 3 shows a CART model, a method that classifies patients according to their likelihood of death using SpO_2_/FiO_2_, Hb, age, and NLR as independent variables. The patients with the highest probability of death (90%) were those who had SpO_2_/FiO_2_ <324, were older than 62 years, and had an Hb >16 g/dL. Additionally, patients with an Hb <12 g/dl and NLR >4 had a higher probability of death (57%). In comparison, patients with the lowest likelihood of death (10%) were those who did have an SpO_2_/FiO_2_ >324 and an Hb >12 g/dL at admission. Interestingly, the CART method also showed that the hemoglobin concentration at admittance modified the SpO_2_/FiO_2_ effect on probability of death.

**Fig. 3.**
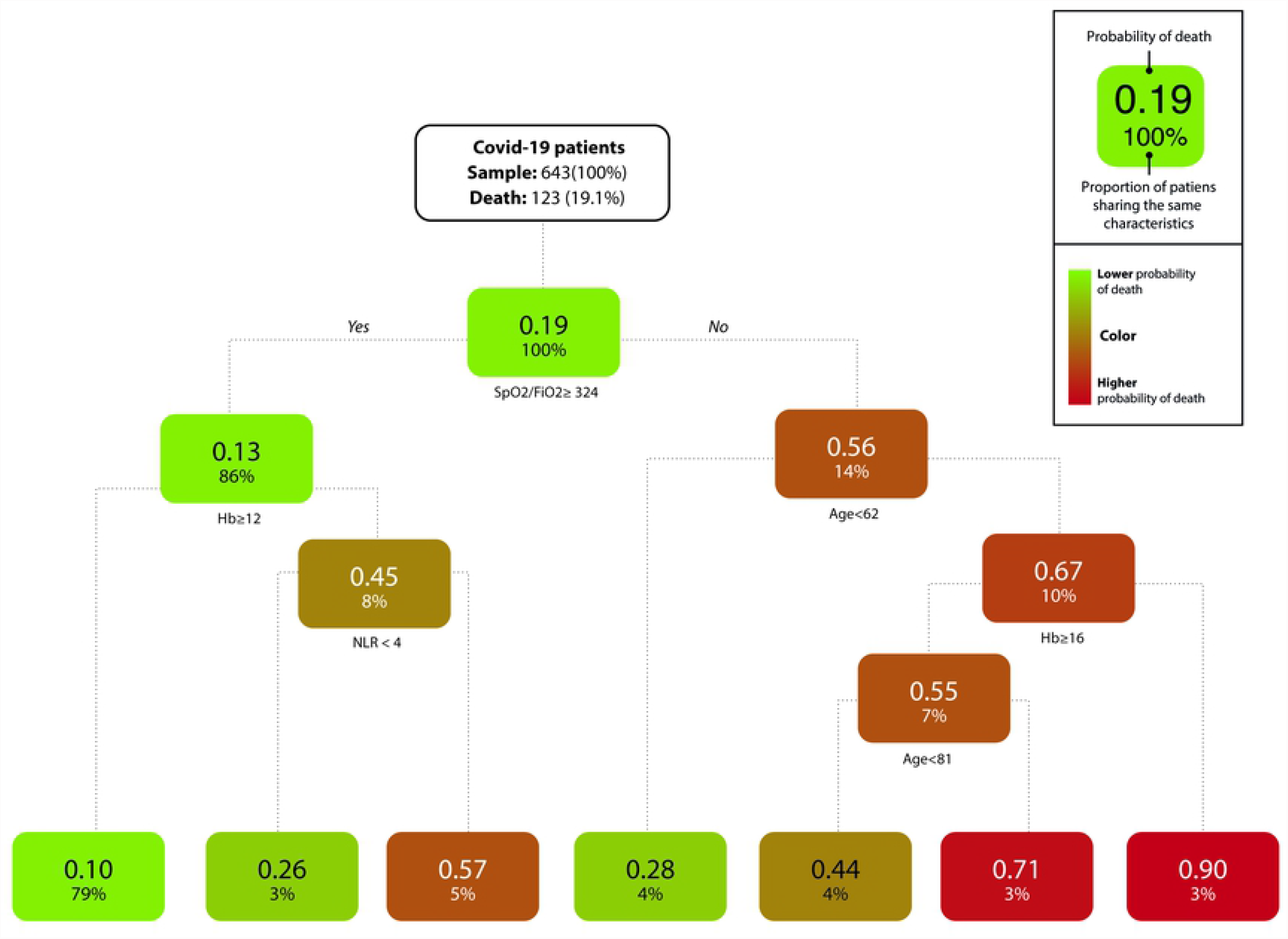
Classification and regression tree of the mortality of COVID-19 patients. **Figure 3**. Distribution of death and survival across COVID-19 patients. The CART showed seven differential clinical profiles using SpO_2_/FiO_2_, age, Hb concentration, and NLR as predictor variables. Clinical profiles with a lower probability of death are shown in green, while red indicates the clinical profiles with a higher likelihood of death. Notice how hemoglobin concentrations both <12 g/dL and ≥16 g/dL are features of the clinical profiles with higher mortality likelihood based upon SpO_2_/FiO_2_ admission value. Hb: Hemoglobin (g/dl), NLR: Neutrophil to lymphocyte ratio.

## Discussion

This original research described the nonlinear relations and, to our knowledge, an unrecognized interaction effect between Spo_2_/FiO_2_ and Hb on the mortality of patients with SARS-CoV-2 acute respiratory infection. Using novel multivariate statistical methods to assess nonlinear relations, we found that Spo_2_/FiO_2_, Hb, NLR, age, history of hypertension, and male sex were independently associated with the probability of death. The Spo_2_/FiO_2_, Hb, and NLR values had a nonlinear relationship with the outcome in GAM. Interestingly, low or high Hb concentrations showed a sharp quadratic increase in the likelihood of death (Fig. 1). We found a significant interaction between SpO_2_/FiO_2_ and Hb effects on mortality (Fig. 2). We explored this further using CART, which showed that older patients with a lower SpO_2_/FiO_2_ and a Hb ≥16 g/dL had the highest probability of death (90%). Additionally, normoxic (SpO_2_/FiO_2_ >324) but anemic patients (<12 g/dl) with an NLR >4 had a higher probability of death (57%). Normoxic nonanemic patients (>12 g/dl) had, as expected, the lowest likelihood of death (10%).

The proportion of death among hospitalized patients was 19.12%, and the cumulative death incidence between March and July was 95.6 per 500 patients admitted to HUM. The Centers for Disease Control and Prevention (CDC) reported in-hospital mortality ranging from 8.6% to 23.4% in approximately the same period.[27] Regarding the oxygenation index, we found a significant difference in the median SpO_2_/FiO_2_ between survivors and nonsurvivors. Some authors have proposed the SpO_2_/FiO_2_ ratio as a continuous noninvasive monitoring tool for assessing oxygenation in acute respiratory failure, or its use as a surrogate for estimating PaO_2_/FiO_2_ in sepsis.[28,29] The SpO_2_/FiO_2_ ratio showed good performance with an AUC =0.801 (95% CI 0.746–0.855) for early mechanical ventilation requirements in patients with COVID-19, even with better performance than the ROX index. Its sequential assessment showed a sharp decline in nonsurvivors compared to an increasing shift in survivors.[30] Catoire et al. described an excellent performance AUC of 0.91 (95% CI 0.885–0.950) of SpO_2_/FiO_2_ to estimate PaO_2_/FiO_2_ values in patients with COVID-19 patients.

We found a significant association between SpO_2_/FiO_2_ and mortality. The GAM showed a sharp increase in the probability of mortality as SpO_2_/FiO_2_ decreased (Fig. 1), with a slight decrease at values under 100. The CART model established a cut point of 324 to define higher-risk groups. Type I respiratory failure in acute respiratory distress syndrome (ARDS) is stratified by PaO_2_/FiO_2_ evaluation, although some authors have proposed a modified definition for scarce resource settings using SpO_2_/FiO_2_ assessment.[31] Berlin’s definition has allowed stratifying patients based on oxygenation compromise. There is a correlation between disease severity measured by PaO_2_/FiO_2_ and mortality risk. However, it has a limited prognostic performance with an AUC of 0.57 (95% CI 0.561-0.593).

For instance, patients with severe ARDS (PaO_2_/FiO_2_ values <100) have a differential prognosis according to distensibility and expiratory volume. Higher expiratory volume and lower distensibility distinguish a higher mortality risk group, even with similar PaO_2_/FiO_2_. [32] Nevertheless, these measures require advanced monitoring, which is not frequently available in scarce resource settings. Numerous studies have explored other biomarkers to enhance prognostication in patients with ARDS.

Red bloodline characteristics have raised concern in the physiopathology of COVID-19 and ARDS. In our study, either high or low Hb values were associated with higher likelihood of death, giving a nonlinear, U-shaped correlation between hemoglobin concentration and probability of death in GAM (Fig. 1C). However, the oxygenation index measured by the SpO_2_/FiO_2_ ratio modified this effect. Patients with similar SpO_2_/FiO_2_ had different mortality probabilities according to Hb concentration (Fig. 2).

Taneri et al. conducted a meta-analysis including 139 observational studies. They reported a weighted mean difference (WMD) of -4.08 g/L (CI -5.12; -305) between moderate and severe cases in patients with COVID-19. However, among 27 studies, there were no significant differences in the pooled mean Hb concentrations between survivors and nonsurvivors, with a WMD of -0.26 g/L (95% CI -2.37; 1.85).[21] In an in silico analysis, Liu et al. proposed a direct effect of SARS-CoV-2 proteins ORF1ab, ORF10, and ORF3a in the Hb beta chain competing against iron. This leads to a functional loss of hemoglobin and hemolysis. However, given that these hypotheses proposed a nondescribed protein–protein hemoglobin degradation pathway, some authors criticized it for its flawed methods and unreliability.[33,34] In the same way, De Martino et al. did not find differences in the hemoglobin dissociation curves or hemolytic biomarkers between patients with COVID-19-related ARDS and patients with ARDS from other causes.[35]

Histopathological descriptions of the lung tissue of patients with ARDS describe the presence of RBCs. However, they are considered a marker for the increased permeability of the endothelial-alveolar barrier. Nevertheless, recently, the role of RBCs and cell-free Hb has been discussed as a central phenomenon in the progression of acute respiratory infection to sepsis, critical illness, and ARDS pathogenesis. The proinflammatory state steers lipid peroxidation, pump damage, changes in calcium influx, and 2,3-DPG concentrations. This leads to RBC membrane changes, facilitating their aggregation, inducing thrombotic events, and hemolysis with cell-free hemoglobin and iron unstable (Fe 4+) group liberation. These perpetuate an injury cycle with nitric oxide consumption, vasoconstriction, inflammation, and increased endothelial permeability.[36] Oxidative and inflammatory damage, and thrombosis with cell aggregation in capillary beds, might explain the harmful effect of polycythemia that we observed in our sample.

Furthermore, other RBC biomarkers are related to ARDS prognosis. Some authors have studied red blood cell distribution width (RDW) and circulating nucleated red blood cells (NRBCs) as prognostic markers. A RDW >14.5 was independently associated with mortality at 30 and 90 days (OR 1.91, CI 95% 1.08–3.39 and 2.56, CI 95% 1.50–4.37, respectively).[37,38] The NRBCs are biomarkers of increased erythropoietic activity. They are related to hypoxemia and inflammation. A study reported a significant difference in the proportion of deaths (50.8% versus 27.3% [p <0.001]) between patients with and without NRBCs. Additionally, patients with severe cases had a higher number of NRBCs, and they were detectable for a longer time. Additionally, there was a negative correlation between the NRBC absolute count and survival time.[39]

Otherwise, we found that a higher NLR was related to worse outcomes. Consistently, previous studies have proposed this relation as a biomarker of severe disease.[40,41] It is used in validated evaluation scales such as COVID-GRAM for predicting critical illness risk.[40] Kilercik et al. evaluated the performance of CBC measures for predicting risk of COVID-19 severity and mortality. They reported that an NLR >5.23 had a sensitivity and specificity for mortality of 85.6% and 56.8%, respectively. Additionally, with a cutoff point of 4.4, NLR had a sensitivity of 78.5% and specificity of 68.2% for predicting severe disease.[42]

A lower ALC is related to the risk of COVID-19 progression. The CALL score uses it as a variable for predicting pneumonia progression.[43] Patients with COVID-19 who developed ARDS demonstrated sustained CD4+ and CD8+ lymphopenia compared to patients without COVID-19-related ARDS.[44] Additionally, the absolute counts of B, T, and natural killer (NK) cells were found to be significantly lower in patients with severe COVID-19 cases than in patients with nonsevere COVID-19 cases.[8] Immune activation by long-term antigen exposure throughout life is related to sterile and chronic low-grade inflammation associated with chronic diseases, cardiovascular risk, obesity, and cancer.[45] This process leads to a senescent adaptive immune system with a predominance of an innate response, which could generate an exaggerated inflammatory response leading to sepsis. The NLR could be a biomarker of this predominance.[44,45]

We also found that male patients were more likely to die in the hospital than females. Analysis of other coronaviruses, such as SARS and MERS, also showed an excess of mortality in male patients. [46,47] Interestingly, the erythroid response to chronic high-altitude hypoxia might be influenced by sex hormones. Chronic mountain sickness (CMS) is rare in Andean females of reproductive age, but its incidence presents a sharp increase with menopause. Additionally, CMS is more frequent in Andean male patients than in female patients. Estrogens might confer protection against excessive erythrocytosis in CMS or Monge’s disease. At physiologic concentrations in vitro, estrogens were related to lower RBC counts than testosterone. Estrogens regulate *EPO, HIF1A, GATA1, VEGF*, genes related to erythropoiesis and erythroid apoptosis mechanisms.[48] These findings suggest that sex hormones moderate the erythropoietic response of chronic exposure to hypoxia and may contribute to the mortality excess in male patients with COVID-19.

This study has some limitations. First, we only considered admission values as predictors of mortality. Patients with COVID-19 might have different behaviors with several kinds of intervention requirements that we did not observe, and which could modify the prognosis. Additionally, we did not consider certain variables related to the Hb dissociation curve as PCO_2_ or acid-base status, which might be associated with residual confusion bias. Furthermore, we did not consider the pharmacological history that could affect the baseline CBC. Lastly, given the retrospective nature of our study, and since the data were gathered from clinical records, we could not guarantee a uniform measurement for each variable. It is necessary to note that no effect can be associated with high-altitude exposure given the absence of a comparative group. The variables studied might also be modified in chronic pulmonary disease or tobacco use. However, including those variables in the purposeful selection of covariates for multivariate analysis showed us that they lacked significance. This study allowed us to hypothesize that the adaptative mechanism for chronic hypoxia modifies the pathological process in respiratory diseases. Further studies are required to understand this biological interaction.

## Conclusion

This study explored the role of the oxygenation index and CBC biomarkers taken at admission as predictors of in-hospital mortality. We found in GAM that SpO_2_/FiO_2_, Hb, and NLR were independently associated with mortality following nonlinear trends. Both low and high Hb concentrations showed a higher likelihood of death. Decreases in SpO_2_/FiO_2_ were associated with a sharp increase in the likelihood of death. However, we found that the effects of Hb and SpO_2_/FiO_2_ on mortality were modified by each other. For instance, patients with similar oxygen index values had different death probabilities based on their Hb at admission. The likelihood of death of patients with a low SpO_2_/FiO_2_ increased proportionally as Hb increased. The CART model showed that patients with a SpO_2_/FiO_2_ >324 and an Hb >12 g/dl had the lowest mortality risk (10%). In contrast, normoxic but anemic patients with Hb <12 g/dl and NLR >4 had a higher probability of death (57%). Finally, patients whose SpO_2_/FiO_2_ was lower than 324, who were older than 62 years, and who had a Hb ≥16 g/dl had the highest mortality risk (90%).

## Data Availability

All relevant data are within the manuscript and its Supporting Information files.

## Author contributions

AMPR and AMRS led and supervised the execution of the research. They contributed to evidence interpretation, critical reviewing and commenting on the manuscript.

DRRL conceptualized the study, conducted the project, collected the data, interpreted the evidence, and critically reviewed the manuscript.

NMG contributed to data curation, formal analysis, and visualization.

AFPA conceptualized the study, worked in data curation, formal analysis, visualization, evidence interpretation, and manuscript drafting.

## Acknowledgments

We thank Estefania Rodríguez Alvarino, Isabella Sanclemente Mariño, and Mateo Díaz Quiroz for supporting the literature review, and Elizabeth Cruz Tapias for her work in image editing.

## Notes

### Abbreviations

AIC: Akaike information criteria
ALC: Absolute lymphocyte count
ANC: Absolute neutrophil count
ARDS: Acute respiratory distress syndrome
BMI: Body mass index
CART: Classification and regression tree
CBC: Complete blood count
CKD: Chronic kidney disease
CMS: Chronic mountain sickness
COPD: Chronic obstructive pulmonary disease
EPO: Erythropoietin
FiO2: Fraction inspired
O2; GAM: Generalized additive model
Hb: Hemoglobin concentration (g/dL)
HIF-1a: Hypoxia-inducible factor 1-α
HIV: Human immunodeficiency virus
HUM: Hospital Universitario Mayor
MAP: Mean arterial pressure
MASL: Meters above sea level
NK: Natural killer cells
NLR: Neutrophil to lymphocyte ratio
NRBCs: Nucleated red blood cells circulating
PCO2: Partial CO2 pressure
RBC: Red blood cells count
RDW: Red blood cell distribution width
SpO2: Peripheral O2 saturation
VEGF: Vascular-endothelial growth factor
WBC: White blood cell count
WMD: Weighted mean difference.

